# Real-world use, safety, and mental health efficacy of regulated psilocybin services in Oregon and Colorado

**DOI:** 10.64898/2026.08.11.26360133

**Authors:** Scott M. Thompson, Devin P. Effinger, Andrew M. Novick, Sara C. Bates, Andrew Scott Conley, Christopher Tobin-Campbell, C. Neill Epperson, Nikolai Rex Skievaski

**Author notes:** To whom correspondence should be addressed at; Department of Psychiatry, 12700 E. 19^th^ Ave., Aurora, CO 80045. Data collected by Althea PBC was analyzed at the University of Colorado School of Medicine.

## Abstract

Oregon (OR) and Colorado (CO) were the first states to enact regulations for provision of psilocybin with support of licensed ‘facilitators.’ As more states and countries adopt similar policies, informed public policy decisions require that client characteristics and rationale for using psilocybin, psilocybin dosing practices, mental health outcomes, and adverse events are understood. We performed a retrospective observational study of responses for 2363 individuals receiving psilocybin at OR and CO regulated service centers. Clients and facilitators entered data before and after receiving psilocybin, including the Mystical Experience Questionnaire-30 (MEQ-30), Patient Health Questionnaire-9 (PHQ-9), Generalized Anxiety Disorder-7 (GAD-7), and World Health Organization Well-Being Index-5 (WHO-5). Preexisting mental health issues were common (66%) in participants. Psilocybin doses ranged from 1-95 mg, with a mean total of 28·8 mg. We observed improvements of 49% in PHQ-9 scores, 51% in GAD-7 scores, and 22% in WHO-5 scores at two-weeks after dosing. MEQ-30 scores were dose-dependent. Changes in PHQ-9 and GAD-7 scores were not different for psilocybin doses ≤30 mg and >30 mg, and only weakly correlated with MEQ-30 scores. There were 94 mild adverse events during and after dosing, five more serious events not clearly related to treatment, and evidence of possible risk of increased suicidality. Study limitations include open label administration, self-reporting, loss of participants for follow-up, and a short 2-week post-dosing end-point. We conclude that psilocybin services, delivered within these regulated frameworks, is associated with improvements in mental health in real world populations, however, more robust monitoring is needed to ensure safety.

## Introduction

Psilocybin, in combination with psychological support, has shown promise in clinical trials for a broad spectrum of psychiatric disorders, including depression,^1^ anxiety,^2^ and substance use disorders.^3^ These improvements are rapid and may persist for 12 months.^4,5^ Treatment-emergent adverse events are generally mild in clinical trials, with careful pre-screening of participants.^6–8^ Whether these results generalize to real-world populations is uncertain but important for the public and policy makers.

Oregon (OR) and Colorado (CO) were the first American states to legalize the regulated administration of psilocybin. Both states use a process that resembles contemporary clinical trial protocols, including mandatory preparatory, administration, and integration sessions. Psilocybin is administered in a licensed ‘service’ (OR) or ‘healing’ (CO) center under the supervision of a licensed ‘facilitator.’ Facilitators may provide any form of supportive or psychologically informed guidance during administration sessions. Mushroom-derived psilocybin products are prepared in state-licensed facilities and tested for purity and potency. Both states require participant safety screening prior to administration, documentation of demographic and dosage information, and reporting of adverse events during and after administration.^9,10^ Participants who screen positive for suicidal ideation (OR) or use of antipsychotic medications (OR and CO) are not allowed to receive psilocybin.

As a service to facilitators and centers, Althea, a Public Benefit Company, created app-based tools to document compliance with state laws. In order to evaluate the efficacy of the regulated psilocybin services, Althea also implemented the voluntary collection of additional pre- and post-dosing information from consenting clients. We analyzed these data to characterize who seeks psilocybin under this regulated pathway, their motivations for seeking psilocybin, and changes in their mental health after dosing. We further documented their dose of psilocybin, the subjective effects they experienced, and the prevalence and nature of adverse events.

## Methods

This is a retrospective observational study of a database collected between December 2024 and March 2026 from clients receiving regulated psilocybin services in CO and OR. All sessions were documented through a single digital platform used by participating facilitators and centers, which applied a common data schema for demographic, dosing, and adverse-event records regardless of program or regulatory jurisdiction. The optional pre- and post-dosing research instruments were also administered through this platform, allowing individual-level outcome data to be linked to standardized dosing and safety records rather than assembled post hoc from heterogeneous sources. The results are reported consistent with STROBE guidelines.

The collection of data was initiated at the preparatory session (**Fig 1**). The participant and facilitator completed the state-mandated screening process and additional questions covering demographics, reasons for seeking psilocybin-assisted therapy, prior psychedelic experience, physical, emotional, and mental health status, suicidality, substance use, and medication use. In Althea asked for voluntary completion of standard self-report instruments: the PHQ-2,^11^ GAD-2,^12^ and the WHO-5 questionnaire.^13^ the Participants scoring ≥3 on the PHQ-2 or GAD-2 were invited to complete the full PHQ-9^14^ and GAD-7^12^, respectively. Prior to submitting these optional responses, participants agreed to Althea’s Terms and Conditions, including a disclosure that de-identified data may be used for research.

**Figure 1.**
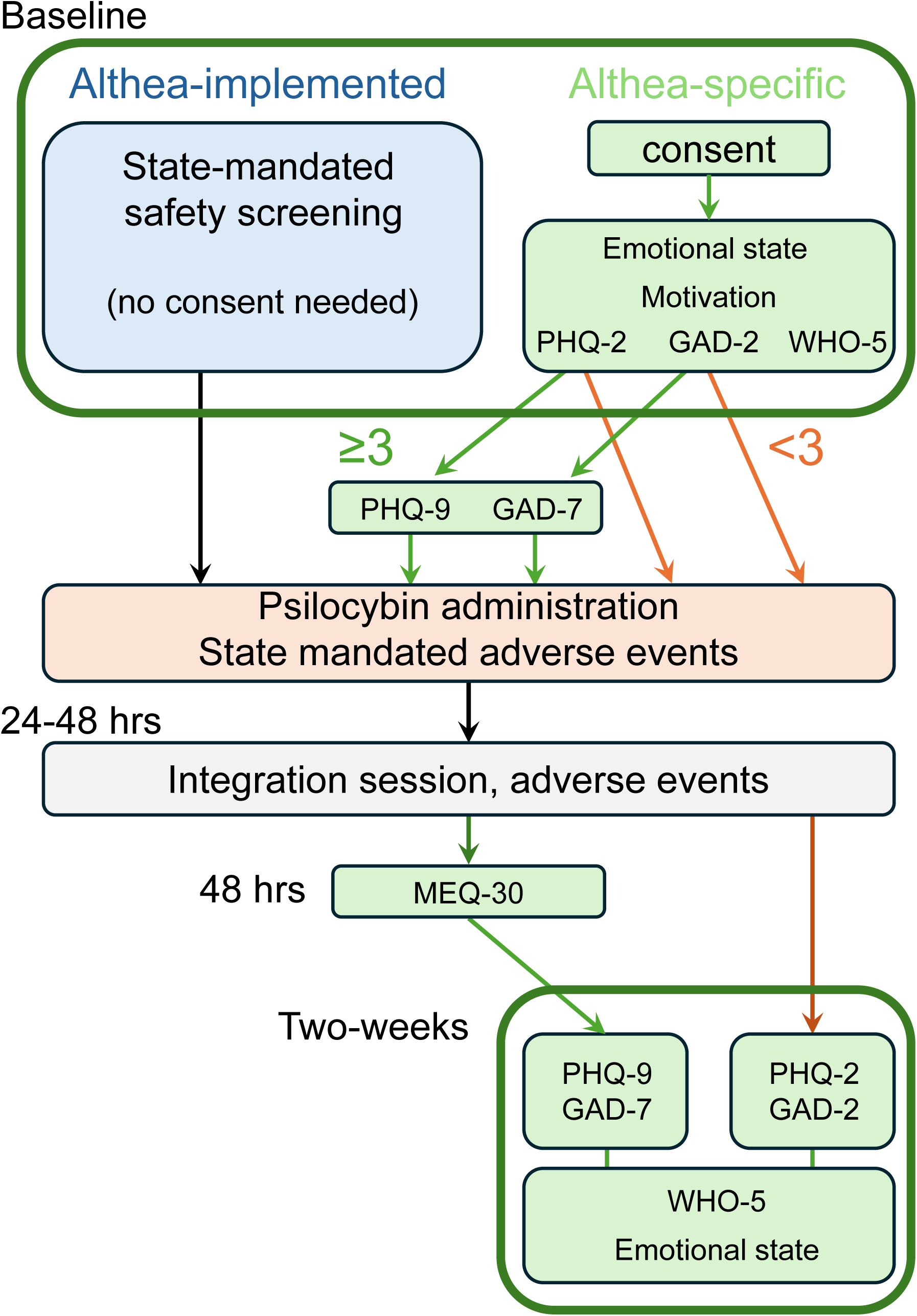
Data collection process flowchart. All participants were required by state law to answer baseline state-mandated safety-related questions. Some participants were also asked to voluntarily complete additional questions at baseline and gave consent to have their de-identified data used for research purposes. These questions included the PHQ-2, GAD-2, and WHO-5, as well as additional questions related to their emotional state and motivation for seeking psilocybin services. Participants whose PHQ-2 or GAD-2 scores were ≥3 then asked to complete the remaining questions from the PHQ-9 and GAD-7. At the dosing session, facilitators recorded the dose(s) of psilocybin administered and any adverse behavioral or mental health events. Participants then completed their integration session and any post-dosing adverse events were also recorded. Participants were asked to complete the MEQ-30 48 hrs after their dosing session. Finally, participants were asked to complete post-dosing the PHQ-2 or -9 and GAD-2 or -7, as well as the WHO-5 and questions about their emotional state. Our data set, which includes 2363 participants who completed only the state-mandated screening questions and participants who also answered the optional questions, was collected by software created and implemented by Althea PBC.

At the dosing session, facilitators entered the dose(s) of psilocybin administered and descriptions of any adverse ‘behavioral’ and ‘medical events,’ as defined by state law.^9,10^ Examination of the data revealed that the labeling between the two categories was not consistent between facilitators, and we therefore combined them in our analyses. Psilocybin was administered in multiple forms, including dried mushrooms, teas, or other edible forms, but each had a potency specified by the licensed manufacturer. Only clients receiving psilocybin at doses of 1 -100 mg are included in the analyses (N= 2363), under the assumption that doses out of this range were mis-entered into the database.

At the integration session, participants were asked to complete the Mystical Experience Questionnaire (MEQ)-30^15^ to characterize their subjective experiences under the influence of psilocybin. Two-weeks after dosing, participants were asked to complete the PHQ-2, GAD-2, and WHO-5 and, if they completed them prior to dosing, the PHQ-9 and GAD-7 again, allowing for pre- and post-dosing comparisons.

Descriptions of any medical and behavioral post-dosing adverse events were collected at the dosing and integration sessions.

At baseline, 1507 participants completed the PHQ-2, GAD-2, and WHO-5. At two weeks post-dosing, 600 participants had paired pre- and post-dosing WHO-5 data, 138 had paired PHQ-9 data, and 181 had paired GAD-7 data. These numbers are less than the number of participants receiving psilocybin because use of Althea’s baseline screening questionnaires was optional, and some facilitators and centers used other screening workflows before documenting dosing sessions in the platform. Furthermore, answering many questions was optional and some participants chose not to respond.

All data were de-identified by Althea and provided to the CU Anschutz researchers for analysis. Numerical data are presented as mean ± STD. The data analysis process was approved by the CU Anschutz Institutional Review Board (protocol # 26-0913).

## Results

### Program demographics

Psilocybin was administered by 253 facilitators. The number of participants treated per facilitator varied from 1 – 123. Of 2363 participants receiving psilocybin, 1954 (82%) were in OR and the remainder in CO.

### Participant demographics

Of 1507 clients who responded to the prompt, 833 (55%) identified as female, 644 (43%) identified as male, 30 (2%) identified as non-binary. The racial and ethnic self-identity of the participants was predominantly White (1228 of 1476 respondents, 83%). The mean age of the participants was 48.1 ± 12.9 years (range=21 – 84).

Residents of CO and OR comprised 44% of participants. The remaining 56% of participants traveled to CO or OR to receive psilocybin services. All 48 other US states were included in this subset of participants, as well as 34 residing outside the US.

Prior psychedelic use was reported by 42% (991) of participants. Of those, 23% (223) reported a prior negative experience. They reported no severe adverse events in their current experience.

Of 1476 respondents, the primary motivations for seeking psilocybin services were trauma, depression, anxiety, and other mental health concerns (531, 36%), personal growth and change of perspective (355, 24%), general health and wellness (296, 20%), expanded consciousness, and spirituality (246, 17%)(**Table 1**). The mean score of 1507 respondents on the PHQ-2 score was 1·67 ± 1·84 and the mean GAD-2 score was 2·14 ± 1·79. Of these 1507 participants, 368 had a PHQ-2 score ≥3 (24%) and 478 had a GAD-2 score ≥3 (32%). 66% of respondents reported a history of depression and/or anxiety (981 of 1476), 82% reported trauma history (730 of 887), and 54% (799 of 1476) were currently seeing a mental health provider, highlighting the prevalence of mental health issues and therapeutic need in the community. Seeking help for substance abuse was not common (30 of 1476, 2%).

**Table 1.**
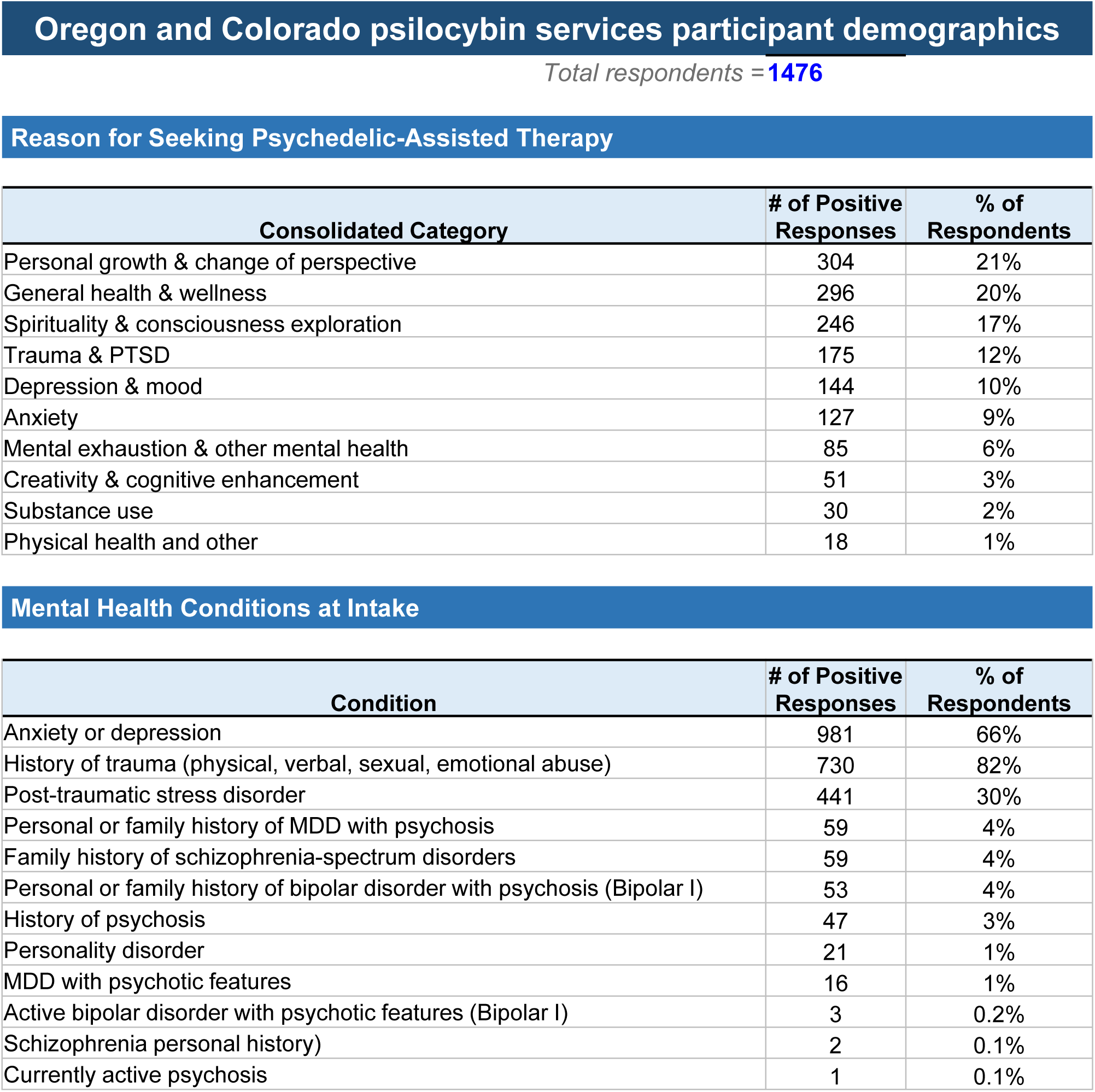
Participant demographics for Oregon and Colorado psilocybin services. TOP: Number and percentage of respondents who endorsed a specific reason for seeking psilocybin services at preparatory session. Respondents were only allowed one selection. Percentages sum to 100%. BOTTOM: Number and percentage of respondents who endorsed having a mental health condition at the time of the preparatory session. Respondents could report multiple conditions. Percentages do not sum to 100%.

### Dosing and subjective effects

The dosing of psilocybin was collected for 2363 sessions. In most sessions, only a single dose of psilocybin was administered, however, in 40% of sessions (1108 of 2755), a smaller second dose was administered within hours the first. We will refer throughout to the total dose a participant received in one or two sessions in a 24-48 hour period. The mean total dose was 28·8 ± 11·6 mg, comparable to the typical 25 mg dose used in clinical trials.^1,6,7^ The range of doses administered was considerable, ranging from 1 mg to 95 mg.

Participant responses to the MEQ-30 were collected at two-weeks post-dosing. As expected, the total MEQ-30 score varied with the total dose of psilocybin administered (**Fig 2A**). There was no effect of gender on the dose-dependence of total MEQ-30 scores (two-way ANOVA, main effect of dose, p<0·0001; no interaction with gender, p=0·18) and no effect of age (<45 vs. ≥45 years of age) (two-way ANOVA, main effect of dose, p<0·0001; no interaction with age, p=0·51). Maximal mean MEQ-30 scores were obtained with psilocybin doses of around 30 mg and did not significantly increase with larger doses (ANOVA: p<0·0001, significant effect of dose; Welch t-test: p>0·05 for all dose bands compared to 20-29 mg band, except <10 mg).

**Figure 2.**
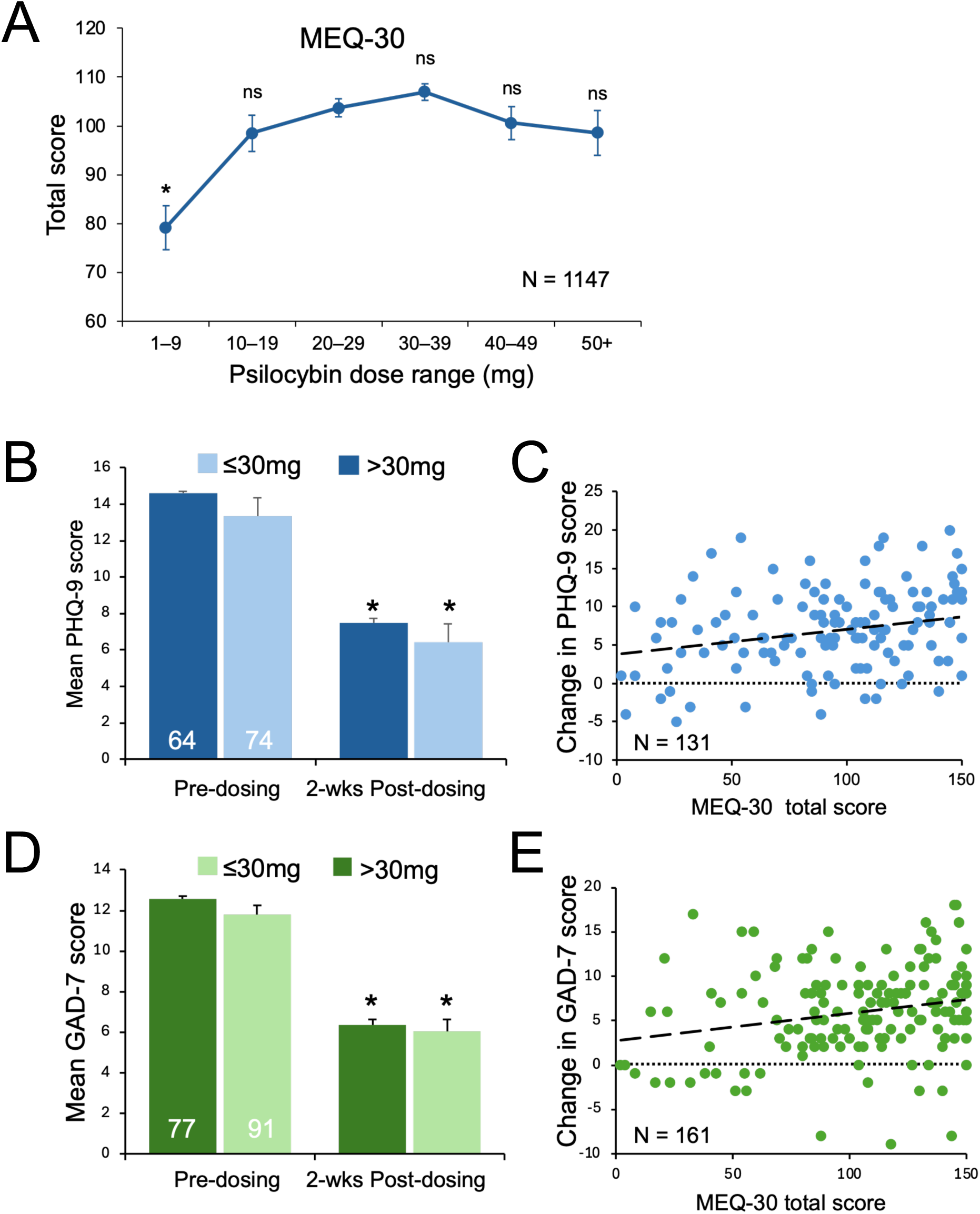
The relationship between psilocybin dose, MEQ-30 score, and mental health outcomes. **A.** Total MEQ-30 score as a function of total psilocybin dose administered. *, p<0·001; ns, not significantly different (p>0·05), Welch’s t-test, compared to mean response to 20-29 mg dose. N, number of participant responses quantified. For the MEQ-30, participants rate 30 items on a 6-point scale, ranging from 0=not at all to 5=extreme. We present only the total score here. The mean and SEM of PHQ-9 (**B**) and GAD-7 (**D**) scores are shown before and two weeks after dosing for participants who received a total dose of psilocybin of ≤30 mg or >30 mg. There was a significant difference for all groups pre- and post-dosing (*, p<0·001, paired t-tests). There was no significant difference between the magnitude of the effects in either group (p=0·22 for PHQ-9, p=0·52 for GAD-7; Welch’s t-test). The pre- to postdosing difference in PHQ-9 (**C**) and GAD-7 (**E**) score for each individual is plotted as a function of their MEQ-30 score and fitted with a linear regression (thick dashed line). There was a correlation for both instruments, but the correlations are weak for both (R^2^=0·05 for both PHQ-9 and GAD-7). The number of respondents is indicated on each graph. Values are mean ± SEM.

### Mental health outcomes

Clinical trials provide evidence that psilocybin-assisted therapy is beneficial for treating depression^1^ and anxiety in people facing life-threatening diseases^2^. We used the PHQ-9 and GAD-7 to assess outcomes in real world participants.

PHQ-9 scores were collected from 138 participants before and two-weeks after dosing. PHQ-9 scores improved by an average of 49% overall (N=138), with 43% improvement in females (n=75) and 57% improvement in males (n=62)(**Fig 3A**). Effect sizes were large (Cohen’s d=-1·13 overall, -0·97 in females, -1·41 in males). Statistically significant improvements were seen regardless of starting PHQ-9 score (**Fig 3B**). Response rates, defined as a ≥50% improvement in score, were 62% for all participants, 56% for females, and 71% for males. Persistence at the 2-week post-dosing timepoint, defined as a pre-dosing score >9 (moderate-to-severe depression) and a post-dosing score <9 (mild-to-minimal), was observed in 53% of participants overall, 43% of females, and 66% of males. Baseline PHQ-9 scores in non-responders vs. responders (p=0·046). Although the dose of psilocybin administered was not different (p=0·17), responders reported a significantly higher MEQ-30 score (p=0·001), specifically for the mystical experience and positive mood dimensions (p<0·0005; p<0·0001), but not for transcendence or ineffability (p=0·41; p=0·12).

**Figure 3.**
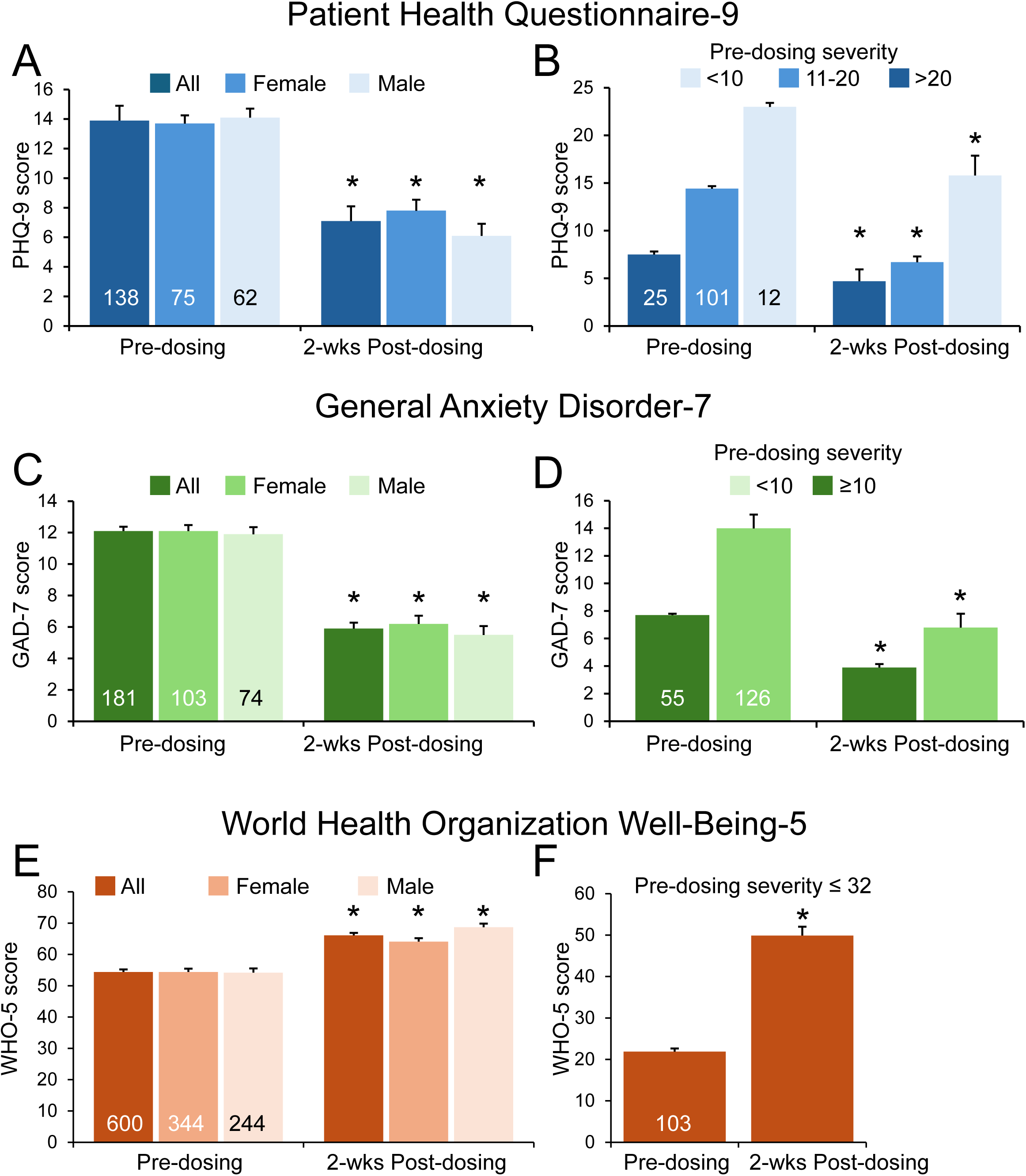
Effects of psilocybin services on self-reported depression, anxiety, and wellbeing. Participant responses to the PHQ-9 (**A,B**), GAD-7 (**C,D**), and WHO-5 (**E,F**) are shown. The mean and SEM scores are shown before and two weeks after receiving psilocybin for all respondents, female, and males in panels **A**, **C**, and **E**. The number of respondents is indicated on the graphs. Responses are illustrated separately for all participants based on the starting severity of their score in **B**, **D**, and **F**. The PHQ-9 is a self-report instrument for depression with a 0-27 point range. Scores of 0-4 are considered to be indicative of minimal or no depression, 5-9 of mild depression, 10-14 of moderate depression, 15-19 of moderately severe depression, and 20 – 27 of severe depression. The GAD-7 is a self-report instrument for generalized anxiety with a 0-21 point range. Scores of 0-4 are considered to be indicative of minimal or no anxiety, 5-9 of mild anxiety, 10-14 of moderate anxiety, 15-21 of severe anxiety. The WHO-5 is a self-report instrument for subjective mental well-being based on responses to five questions, with 0 indicating the worst possible quality of mental health and 5 indicating the best possible quality, with a maximum=25. We present raw scores for the PHQ-9 and GAD-7, and the percentage of the maximum for the WHO-5. Significant pre-post-dosing differences: *, p<0·001, paired t-tests. Values are mean ± SEM.

Pre- and post-dosing GAD-7 scores were collected from 181 participants. GAD-7 scores improved by an average of 51% overall (n=181), with 49% improvement in females (n=103) and 54% improvement in males (n=74)(**Fig 3C**). These sex differences were statistically significant (p=0·036). Effect sizes were large (Cohen’s d=-1·23 overall, -1·15 in females, -1·37 in males). Statistically significant improvements were seen regardless of starting GAD-7 score (**Fig 3D**). Response rates, defined as a ≥50% improvement in score, were 62% for all participants, females, and males. Persistence at the 2-week post-dosing timepoint, defined as a pre-dosing score >5 (mild-to-severe) and a post-dosing score <5 (minimal), was observed in 45% of respondents overall, 43% of females, and 50% of males. There was no significant difference in baseline GAD-7 score (p=0·84) or dose of psilocybin administered (p=0·10) between responders and non-responders, although responders reported higher MEQ-30 scores (p=0·002).

We next asked whether mental health improvements were correlated with the dose of psilocybin administered. We found that improvements in PHQ-9 and GAD-7 scores were not significantly different for those participants receiving a dose of 30 mg or less compared to those receiving >30 mg of psilocybin (**Fig 2B,D**). Overall, 1422 (60%) of 2363 participants received >30 mg of psilocybin. We next tested the correlations between an individual’s change in PHQ-9 or GAD-7 score and their self-reported MEQ-30 score. While these correlations were positive, the linear correlation was weak, with R^2^ values of only 0·05 for each (n=131, 161, respectively) (**Fig 2C,E**). The MEQ-30 score was not significantly different for those receiving ≤30 mg and >30 mg for either instrument (p=0·66 for both, respectively, Welch’s t-test).

PHQ-9 improvements were greater in participants aged under 45 (56% improvement, n=72) than those 45 and older (41%, n=66)(p=0·047), despite their baseline scores and psilocybin doses (26·3 vs 25·8 mg) not being significantly different (p=0·63 and 0·27). GAD-7 improvements did not differ by age group (55% vs 46%)(p=0·11).

Effects of psilocybin service on well-being were assessed from pre- and post-dosing WHO-5 scores (n=600). WHO-5 scores improved by an average of 22% overall, with a 18% improvement in females (n=344) and 27% improvement in males (n=244)(**Fig 3E**). These sex differences were statistically significant (p=0·005). Effect sizes were robust (Cohen’s d=0·57 overall, 0·47 in females, 0·73 in males). WHO-5 scores increased from 21·8 ± 7·9% to 51·1 ± 21·6% (p<0.0001, n=98) in participants who had poor well-being scores prior to receiving psilocybin services (score ≤32%)(**Fig 3F**).

A total of 211 people participated in two psilocybin dosing sessions within a 48 hr time period. To determine whether repeated dosing produced different mental health benefits, we compared PHQ-9, GAD-7, and WHO-5 outcomes between single-session participants and those who participated in a repeat session. Baseline scores were not significantly different between groups. There were no significant differences in the magnitude of improvement in PHQ-9 (p=0·80), GAD-7 (p=0·75), or WHO-5 (p=0·51) between single and repeat-dose participants, or in their total MEQ-30 (p=0.98)(Welch’s t-test for all), indicating that repeat sessions provide no additional mental health benefits.

Participants reported significant reductions in negative states (worry, anxiety, anhedonia) at the 2-week post-dosing time point, consistent with the PHQ-9, GAD-7, and WHO-5 findings (**Fig 4**).

**Figure 4.**
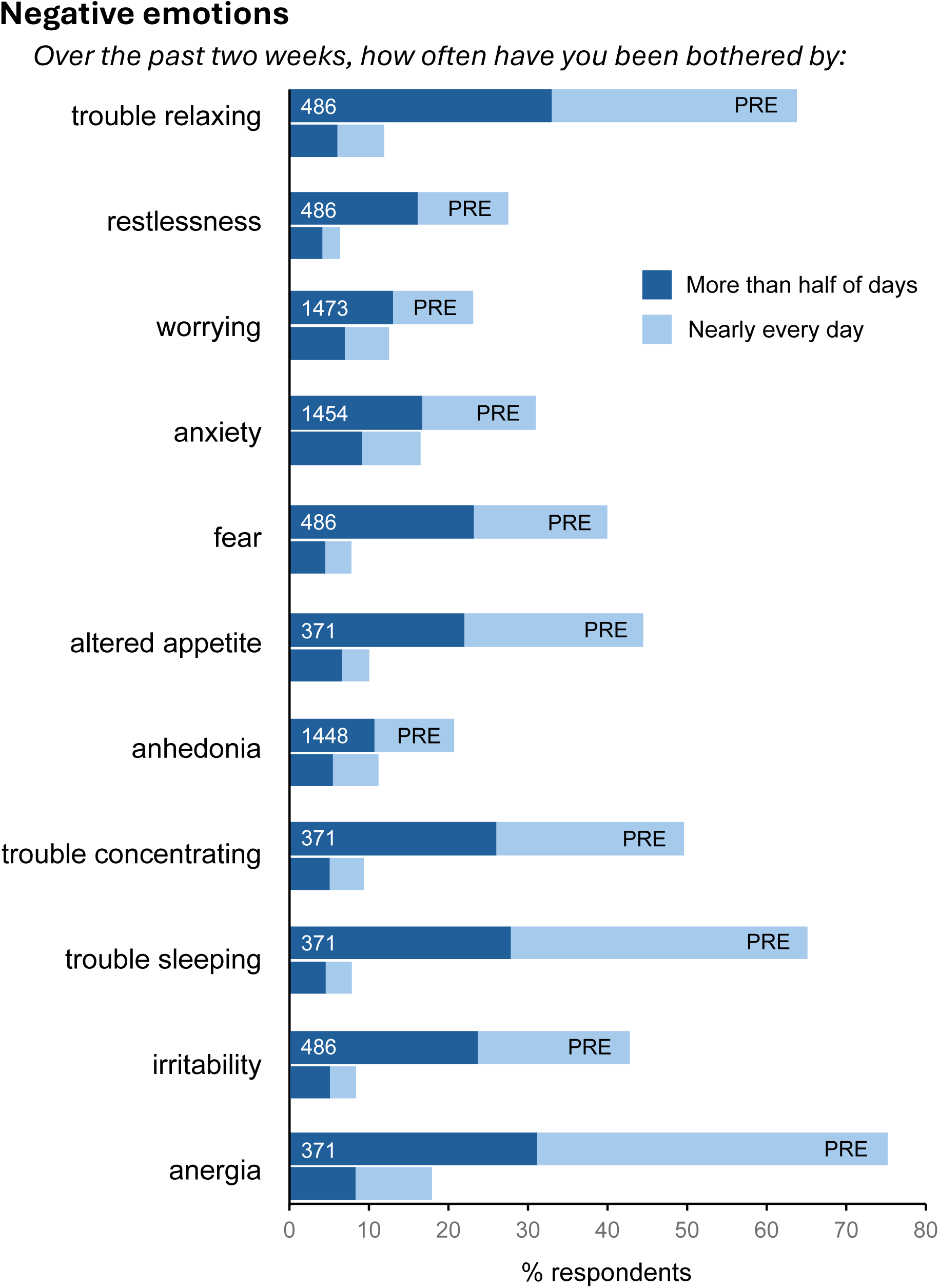
Effects of psilocybin services on emotional health. Respondents were asked a series of questions about negative aspects of their emotional status. The graphs show the percentage of respondents who endorsed “nearly every day” or “more than half of days” in response to concerns over the two weeks before (upper line) and after (lower line) receiving psilocybin. The number of respondents to the pre-dosing questions are shown in the upper bar. The number varies because some questions are taken from the GAD-2 and PHQ-2, and were asked of all participants, some are from the GAD-7 and PHQ9 and were asked of fewer participants. In response to all questions, there was substantial improvement after psilocybin.

### Influence of concomitant SSRI/SNRI use

142 participants reported using SSRI/SNRIs at the time of dosing. These participants received a significantly higher total psilocybin dose than non-users (34·0 vs 27·6 mg, p<0·0001) yet had lower average MEQ-30 scores (96·2 vs 104·1, p=0·022), consistent with a hypothesized downregulation of 5HT2A receptors.^16^ Despite lower MEQ-30 scores, SSRI/SNRI use did not significantly affect PHQ-9 changes (p=0·84) or response rates (p=0·92), or GAD-7 (p=0·41) changes.

### Psychosis

People with a personal or family history of psychosis-related illnesses are almost always excluded from academic and commercial psychedelic drug trials. 59 participants reported personal history of MDD with psychosis, 59 reported a family history of schizophrenia-spectrum disorders, 53 reported a family history of bipolar disorder with psychosis, and 47 reported a personal history of psychosis (**Table 1**). Despite CO and OR screening recommendations, all of these participants received psilocybin, with only three mild adverse events reported (headache, nausea). No new incidents of psychosis-like behavior were reported.

48 participants reported using antipsychotic medications and two reported current treatment with lithium. All received psilocybin in both CO and OR in apparent contravention of their regulations. The mean MEQ-30 for 22 of these participants for whom there was a score was 84·3 ± 46·4 in response to a dose of 36·3 ± 11·7 mg of psilocybin, only slightly lower than responses in those not taking antipsychotics (**Fig. 2A**), suggesting that they may not have been taking antipsychotics at the time of their dosing. Only one mild adverse event was reported for these participants.

### Adverse events

The most common dosing-day adverse events were nausea (35), anxiety (14), crying (6), and agitation (6) across 2363 dosing sessions. Post-dosing adverse events included headaches (16), nausea (4), and memory/confusion (4). No other adverse event received more than 2 mentions pre- or post-dosing. All were reported as mild and transient. There were five more serious adverse events. Two calls for EMS service were made: one for a suspected cardiac event and one for abdominal pain (ultimately determined to be kidney stones). One client requested EMS for emotional distress, but it is not clear if they were called. One client reported several days after dosing that she had had a “heart attack” on dosing day. There was no follow up on any of these events. There was one fall on dosing day without reported injuries.

### Suicide

Of 1476 respondents at baseline, 18 participants endorsed self-harm concerns, 44 had thoughts of suicide, and 87 reported a prior suicide attempt. All received psilocybin. Adverse events for these participants were four reports of nausea on the dosing day, one report of crying on dosing day, and one report of nausea the day after dosing.

We also analyzed responses to the PHQ-9 regarding suicidal ideation and considered responses of *nearly every day, more than half of days, or several days* to be indicative of suicidal risk. Of 371 respondents, 95 expressed suicidal risk in the two weeks preceding their baseline assessment. 28 of these 95 participants responded before and after dosing, of which 22 reported a decreased frequency of suicidal ideation after dosing and 6 reported no change. Of 103 participants who reported no suicidal ideation at baseline, 11 did report suicidal risk two weeks after dosing. Total PHQ-9 scores were increased or unchanged in eight of these 11 after dosing.

### Satisfaction

Asked to rate their experience on a scale of 1 to 10, respondents rated realization of their goals as 7·2 ± 2·3 (n=693), meaningfulness of their experience as 8·7 ± 2·0 (n=694), 9·5 ± 1·1 on the safety of their experience (n=684), and 8·3 ± 1·8 on the benefit of their experience (n=638).

## Discussion

This study analyzes the demographics, motivations, mental health outcomes, and safety of the delivery of psilocybin services in CO and OR under regulations governing licensing of facilitators and healing/service centers, product testing, participant pre-screening, and the recording of adverse events. This is important because creation of similar programs is growing world-wide. Safety and efficacy of psilocybin administration in clinical trials, in which subjects are rigorously screened prior to enrollment, may lack generalizability to real world populations and conditions. Unlike naturalistic or ‘underground’ administration, psilocybin doses are known and standardized procedures were followed in the regulated pathway.

### Demographics

Our sample size is large (2363 participants), comparably distributed between males and females, and spans the adult age spectrum. A significant limitation is that non-White participants represent <20% of respondents.^17^ A majority of participants traveled across state lines,^18^ indicating that availability of these services is shaped in part by the ability to travel, not by need alone.

Mental health concerns were a substantial self-reported motivation for seeking psilocybin services (36%), with 25% having PHQ-2 and GAD-2 scores ≥3, indicative of potential clinically relevant depression or anxiety.^11,12,19^ Half of respondents had prior personal or family mental health issues and were under the care of a mental health provider. Nevertheless, personal growth, wellness, spirituality, and expanded consciousness will also important motivations for seeking psilocybin services, representing 17-24% of all respondents.

### Dosing

The mean dose of mushroom-derived psilocybin administered (29 mg) was comparable to synthetic psilocybin doses used in clinical trials (typically 25 mg) and was sufficient to produce near-maximal MEQ-30 scores. Mushroom-derived products contain other psychoactive components, but these data are consistent with findings on synthetic psilocybin for both peak effective dose (ca. 25 mg) and MEQ-30 score (ca. 115).^20^

Decreased depression and anxiety after psilocybin-assisted therapy. Our data indicate that regulated psilocybin services are comparably effective at treating mental health in the short-term under real world conditions as psilocybin administration in clinical trials.

We observed decreases in PHQ-9 scores of ∼50%, corresponding to improvement from moderate to mild-to-minimal severity. Furthermore, 62% of respondents reported a ≥50% reduction in PHQ-9 score, with robust effect sizes. Improvements were greater in participants under 45 than those 45 and older, despite comparable baseline scores and psilocybin doses, possibly reflecting age-related differences in neuroplasticity.

GAD-7 scores decreased 51%, with 62% achieving ≥50% improvement, consistent across both sexes and baseline severity. These improvements were independent of starting GAD-7 scores, gender, and age.

In parallel with changes in PHQ-9 and GAD-7 scores, participants reported improvements in positive emotional aspects of mood and decreases in negative emotions. Taken together, these diverse self-reported measures paint a picture of significant and broad improvement in mental health across domains.

SSRI/SNRI use complicates psilocybin services because weaning participants is time consuming and potentially dangerous. We observed that SSRI/SNRI use reduced MEQ-30 scores for equivalent doses of psilocybin, consistent with the hypothesized down regulation of 5HT2A receptors, as reported for naturalistic data sets^21^ but not clinical trial data.^22^ Nevertheless, decreases in PHQ-9 or GAD-7 scores were equivalent in SSRI/SNRI users and non-users, suggesting that the ensemble of receptor(s) activated by psilocybin that underlie the therapeutic benefits remain available at sufficient numbers to mediate improvements in depression and anxiety.

Improvements of PHQ-9 and GAD-7 scores were not significantly different for total doses of psilocybin more than or less than 30 mg. Furthermore, equivalent mean MEQ-30 scores were reported by those receiving more than or less than 30 mg of psilocybin. Similarly, we observed only weak correlations between an individual’s MEQ-30 score and improvements in their PHQ-9 and GAD-7 scores (r^2^=0·05), lower than those reported in life-threatening cancer patients (HADS r^2^=0·12; HAM-A r^2^=0·35).^23^ Taken together, we suggest that therapeutic benefits of psilocybin may not occur in a dose-dependent, more-is-better continuum, but perhaps reflect a threshold of some pharmacological and/or psychological response that is necessary and sufficient for inducing a therapeutic response. We further suggest that facilitators and centers need not recommend high doses of psilocybin for their clients because of fears that a lower dose will be insufficient to produce benefits.

Adverse events and psilocybin assisted therapy. Although reporting of adverse events is required by law in CO and OR, the information in this dataset is limited by lack of detail and follow-up.

Nevertheless, contrary to concerns that regulated services would generate high rates of adverse events, the most common reported adverse events were mild and transient (nausea, anxiety, agitation), no worse than in clinical trials,^8^ with no evidence of additional EMS burden. A similar conclusion was drawn recently from analysis of data reported to Oregon regulatory authorities.^24^

Although a recent clinical trial found that psilocybin with psychological support led to improvements in chronic suicidality,^25^ other analyses are more ambiguous.^26^ We observed improvements in self-reported suicide measures in participants with previous suicidal ideation. Although the sample size was small, we also found evidence, however, of de novo suicidal thinking in several non-suicidal participants after treatment. Given the risks, we recommend that monitoring of adverse events, particularly suicidality, be considerably strengthened.

A considerable number of people whose histories of schizophrenia and other psychoses would have excluded them from clinical psilocybin trials received regulated psilocybin services without evidence of adverse consequences. Similarly, the data set contained evidence that participants who may have been using antipsychotic medications and lithium were offered psilocybin services, despite the state regulations. It is uncertain whether they weaned off their medications prior to dosing, however. Reinforcement of the risks and requirements with facilitators would be prudent.

### Limitations

Key limitations of our study include several sources of potential bias. Regulated psilocybin services are provided open-label, leading to placebo and expectancy effects, as in clinical psilocybin trials.^27^ Reliance on self-report instruments may result in reporting inaccuracy and bias.^28^ Furthermore, selective dropout between pre- and post-dosing surveys was apparent, potentially biasing results toward positive outcomes. There are differences in the training and experience of the facilitators, and the extent of the psychological support they provide.^29^ The dataset lacks information on important outcomes like PTSD and SUDs. Lastly, the limited 2-week endpoint does not address longer-term durability of mental health benefits. Nevertheless, these data have a high degree of certainty about the dose and potency of the psilocybin product consumed and the lack of confounding substances that may modulate the effects of psilocybin, such as cannabis and alcohol,^30^ compared to surveys of naturalistic use^31–33^.

Final conclusions. With these limitations, our data provide evidence that regulated psilocybin services in CO and OR can effectively reduce self-perceived mental health challenges in the real world. The data also provide evidence that while psilocybin services are generally safe, risk for suicidality is a significant concern. More broadly, real-world evidence at this scale and consistency depends on standardized data infrastructure embedded at the point of care. The uniform digital capture used here, spanning many independent facilitators and two regulatory frameworks, is what allowed individual-level demographic, dosing, outcome, and adverse-event data to be assembled into an analyzable cohort. Comparable infrastructure should be a precondition as additional jurisdictions adopt regulated psilocybin services.

## Data Availability

Aggregated, de-identified summary data from this ongoing collection are publicly available at https://be.withalthea.com/psilocybin-outcomes. Individual-level de-identified data are not publicly available, owing to the conditions of participant consent and the platform's terms governing research use, but may be made available to qualified researchers on reasonable request to the corresponding author, subject to a data use agreement.

## Author contributions

SMT, conceived data collection process, analyzed data, drafted and edited manuscript, prepared figures. DPE, analyzed data, performed AI tasks, edited manuscript. AMN, provided insight on psychiatric self-report instruments, edited manuscript. SCB, implemented the data collection instruments. ASC, recruited and on-boarded the participating facilitators and centers. CTC, developed the data collection software platform. CNE, provided funding for CU Anschutz team, edited manuscript. NRS, led the Althea team and data collection effort, contributed to project administration, and edited the manuscript.

Use of artificial intelligence. Artificial intelligence (AI) tools were used to assist with Data management and filtering, application of inclusion criteria, and identification of repeat-session participants. Statistical analyses, including paired t-tests, Welch’s independent t-tests, Mann– Whitney U tests, Cohen’s d effect-size calculations, and paired and unpaired baseline-to-post-dosing comparisons, were computed using Python (scipy, pandas) with AI assistance. All statistical results, data values, and citations were reviewed and verified by the authors. The authors take full responsibility for the accuracy and integrity of the reported findings.

The corresponding author affirms that he had access to all de-identified data from the study, both what is reported and what is unreported, and also that he had complete freedom to direct its analysis and its reporting, without influence from the sponsors. The corresponding author also affirms that there was no editorial direction or censorship from the sponsors.

## Data sharing

Aggregated, de-identified summary data from this ongoing collection are publicly available at https://be.withalthea.com/psilocybin-outcomes. Individual-level de-identified data are not publicly available, owing to the conditions of participant consent and the platform’s terms governing research use, but may be made available to qualified researchers on reasonable request to the corresponding author, subject to a data use agreement. The public dashboard is maintained by Althea PBC and updates as data accrue, figures may differ from those reported here, which reflect the December 2024 to March 2026 analysis window.

